# Development of Multiplexed RT-LAMP for Detection of SARS-CoV-2 and Influenza Viral RNA

**DOI:** 10.1101/2020.10.26.20219972

**Authors:** Yinhua Zhang, Nathan A. Tanner

## Abstract

The ongoing COVID-19 pandemic has demonstrated the utility of widespread molecular testing for surveillance and diagnostic detection of SARS-CoV-2. Reverse-transcription loop-mediated isothermal amplification (RT-LAMP) has enabled testing outside of the standard clinical laboratory PCR infrastructure, with simple and rapid tests supplementing the existing, standard methods. However, current LAMP tests have detected single targets and required separate reactions for controls or multiple targets. As flu season arrives in the Northern Hemisphere the ability to screen for multiple viral targets will be increasingly important, and the ability to include internal control assays in the RT-LAMP test allows for decreased resource use and increased throughput. Here we describe a multiplexing approach to RT-LAMP with four targets (SARS-CoV-2, Influenza A, Influenza B, and internal control human RNA) in a single reaction using real-time and endpoint fluorescence detection. This increase to the functionality of RT-LAMP will, we hope, enable even broader adoption of this power molecular testing approach to aid in the global fight against this continuing public health threat.

## Introduction

Reverse-transcription loop-mediated isothermal amplification (RT-LAMP) was developed as a nucleic acid amplification technique requiring only a single reaction temperature without the need for sophisticated thermal cycling equipment [1]. It is particularly well-suited for simple molecular diagnostics as it is rapid and sensitive, with numerous and versatile detection methods for determining positive or negative reactions[2]. In the recent COVID-19 pandemic, LAMP has been successfully applied for SARS-CoV-2 detection using: colorimetric readouts in both simple, point-of-need assays as well as higher-throughput diagnostic platforms [3-11]; Cas enzyme detection for lateral flow or fluorescence [12-14]; and next-generation sequencing, offering high levels of sample pooling and multiplexing [15, 16]. Many of these tests have received Emergency Use Authorization from the FDA or have been CE-marked in Europe, and are used daily for COVID testing worldwide.

However, these initial LAMP tests are generally used for the detection of a single target per reaction, with samples split to multiple reactions for confirmatory detection or controls. A beneficial feature of many RT-qPCR assays, particularly in the widely-used TaqMan® method, is the ability to perform multiplex detection, where tests for numerous analyte targets and internal controls are conducted simultaneously in one reaction. In addition to reducing reagent usage and increasing testing throughput, multiplexing gives a reliable internal control, indicating successful amplification as well as validating sample extraction. Detecting multiple targets from the same pathogen of interest (e.g. Gene N and Gene E from SARS-CoV-2) increases confidence when calling a sample positive, or multiple different targets can be combined for more useful diagnostic tests (e.g. SARS-CoV-2 and influenza). A number of methods exist for multiplexing LAMP, and we previously presented a scheme called Detection of Amplification by Releasing of Quenching (DARQ) that supplements a standard LAMP primer set with a pair of oligonucleotides in duplex form for detection. This duplex consists of a 5′-modified version of the FIP primer (Q-FIP) annealed to a 3′-modified oligo represent the F1 region (Fd), complementary to the FIP in its 5′ section [17]. The two modifications are selected to be dark quencher-fluorophore pairs, and when the quenched FIP is incorporated into the LAMP product, subsequent amplification displaces the Fd oligo, releasing quenching and producing fluorescent signal specific to the label and target of interest. Other versions of this approach have been used, moving the quencher and fluorophore to a Loop primer[18, 19] or incorporated into the amplification products and detected after the reaction [20].

As the COVID-19 pandemic continues into flu season, the ability to distinguish which causative agent is responsible for what can manifest as very similar symptoms will be of great importance for diagnostics and disease surveillance. A patient presenting with respiratory symptoms may have SARS-CoV-2, Influenza A or B, RSV, a rhinovirus, etc. and using one test to identify multiple infectious agents in the same procedure will save time and cost. Most importantly, it gives a more definitive diagnostic identification. Syndromic panel tests (e.g. Genmark, Biofire) are the ultimate example of this principle, with dozens of respiratory pathogens tested for simultaneously but these tests are expensive and not suited to the expanded use of molecular testing that has arisen during COVID. Here, we demonstrate a simple LAMP assay for detection of 4 targets (SARS-CoV-2, Influenza A, Influenza B, and an internal control) together with high sensitivity in a single reaction, expanding the utility of the widely-used LAMP chemistry to a multiplex diagnostic setting.

## Results

We had previously established sensitive LAMP primer sets for SARS-CoV-2 detection [11] and synthesized a DARQ-compatible labeled FIP and Fd oligo (JOE), as well as our internal control primer set (*ACTB*, ROX). For influenza detection, we designed a variety of LAMP primer sets and pulled existing primers from literature reports [21-25] and evaluated them for speed, sensitivity and compatibility with our SARS-CoV-2 and control primers. From this comparison we selected the IAV [21, 25] and IBV [25] primer sets as they were among the most sensitive primer sets in our screening and were designed for suitability with various strains. The IAV set has been validated to detect many Influenza A strains from H1N1 to H15N8 from birds and humans including multiple H1N1/2009pdm and H7N9 strains. In our testing we found it amplified well with RNAs from the 2009 H1N1 (H1N1/2009pdm, ATCC VR-1737D) and 1938 H1N1 (A/PR/8/34, ATCC VR-1469DQ) strains as well as 2012 H3N2 (A/Virginia/ATCC6/2012; ATCC VR-1811D) and 1968 H3N2 (A/Aichi/2/68, ATCC VR-1680D). The IBV set was tested with RNA from a B virus [25] isolated in 2012 and we found it amplified well with RNA from strain B/Wisconsin/1/2010 BX-41A (ATCC-1885DQ), but not from B/Lee/40 strains isolated 80 years ago. We also performed sequence alignments of IAV and IBV target regions and these primer sets are expected to detect strains isolated in recent years. With this information, IAV and IBV primer sets are were chosen for multiplexing with our SARS-CoV-2 primer set using Cy5 and FAM fluorophores, respectively.

First, we compared DARQ LAMP with conventional intercalating dye detection of various concentrations of SARS-CoV-2 RNA using the E1 primer set, either with or without *ACTB* control primer set (Figure 1). The speed by DARQ LAMP showed a target dosage response, just as in the conventional LAMP monitored by SYTO-9 double-stranded DNA binding dye (Figure 1A and 1B). Reaction speed decreased in the presence of the DARQ duplex, and slowed slightly more with addition of a second primer set (*ACTB;* Figure 1C, E). Similarly, in the duplex LAMP reactions *ACTB* amplified slightly slower than reactions containing only *ACTB* and Jurkat RNA (Figure 1D), but the signal level from *ACTB* was sufficient to serve as an internal positive and loading control. Using 0.25X primer concentration for the *ACTB* internal control allowed for even amplification in single- vs. duplex reactions while still providing sufficient fluorescence signal for detection.

**Figure 1.**
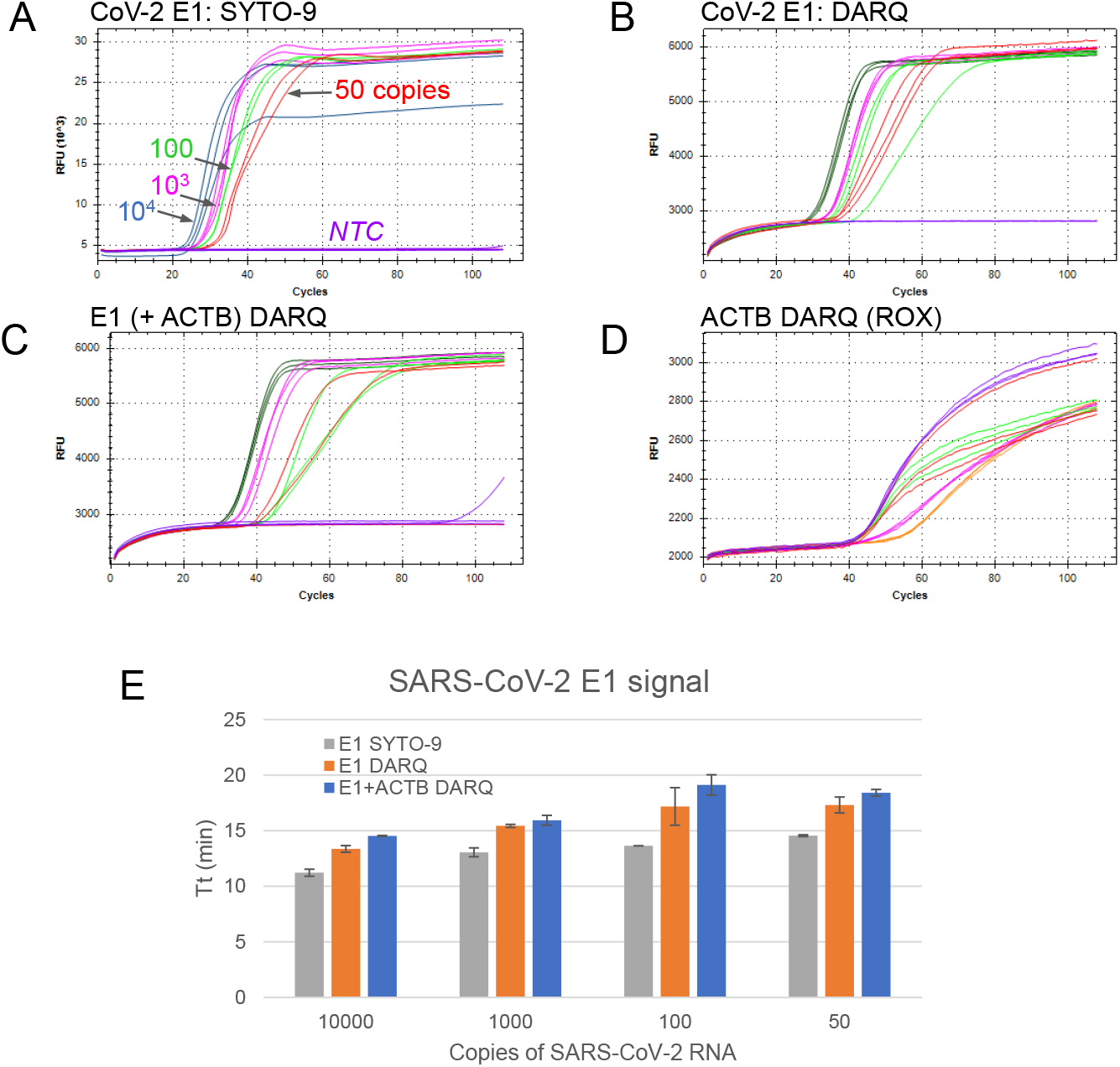
Comparison between conventional and DARQ LAMP. All reactions were performed using E1 primer targeting SARS-CoV-2 RNA at 10^4^, 10^3^ 10^2^ and 50 copies, and NTC in triplicate. A. Conventional LAMP monitored by SYTO-9. B. Single-plex DARQ LAMP. C. Duplex DARQ LAMP including both E1 and *ACTB* primers. D. *ACTB* detection from duplex DARQ LAMP. E. Comparison of SARS-CoV-2 amplification speed in reactions with SYTO-9, single- and duplex DARQ LAMP.

Next, we sought to determine whether DARQ LAMP has similar detection sensitivity as standard LAMP. Figure 2 shows results of 24 LAMP reactions each with 50 copies of SARS-CoV-2 RNA and 8 no-template controls. In all configurations (standard intercalating LAMP (2A); single-plex DARQ LAMP (2B); duplex E1+*ACTB* DARQ LAMP (2C)) we measured a similar number of positive LAMP reactions and negative no-template controls. This indicates that DARQ LAMP has similar detection sensitivity as conventional LAMP reactions, and addition of a second primer set does not increase the rate of non-template amplification with our reaction conditions.

**Figure 2.**
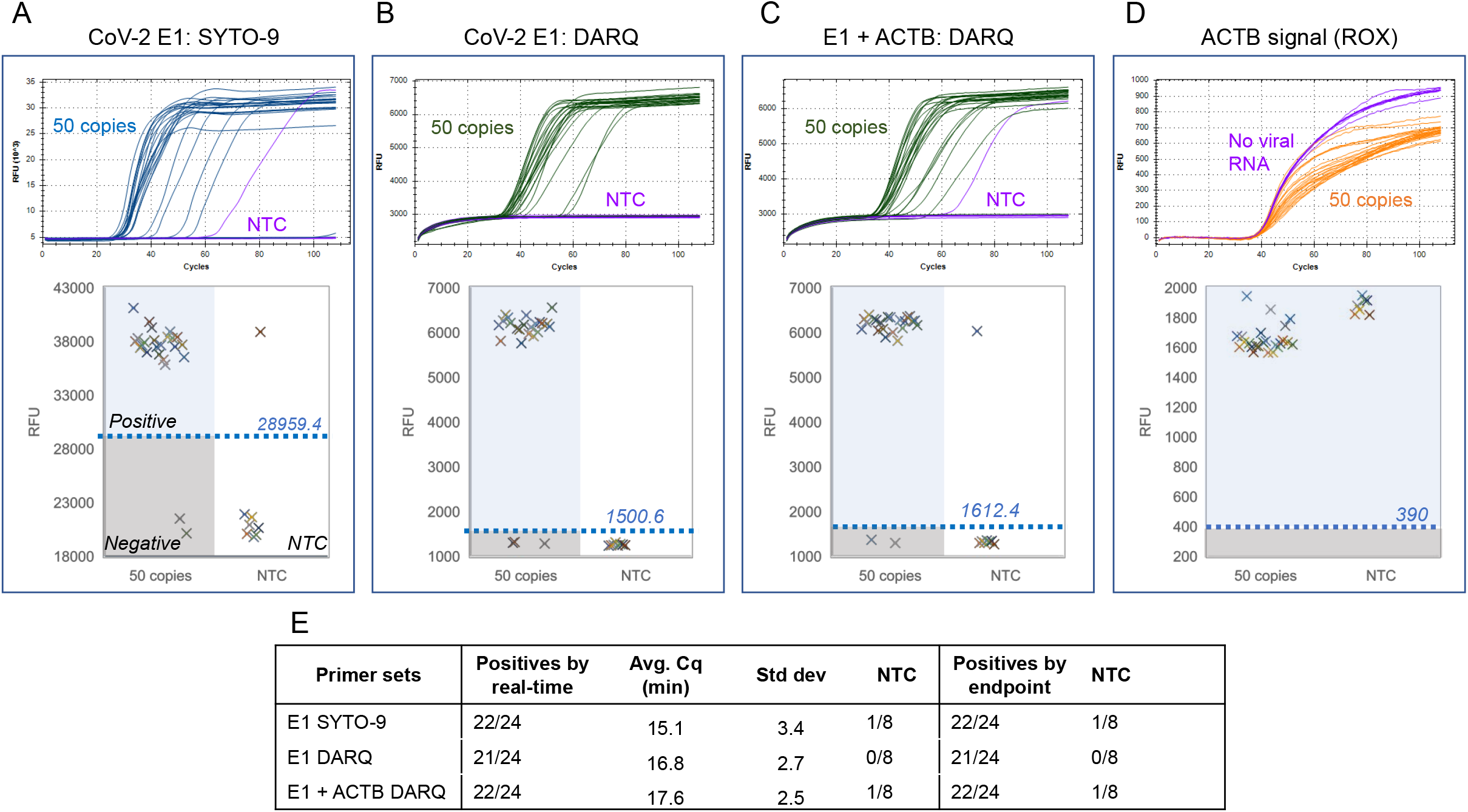
Maintaining detection sensitivity in DARQ LAMP. Amplifications were performed with 24 repeats each containing 50 copies of SARS-CoV-2 RNA or 8 repeats of NTC using E1 or both E1 and *ACTB* primer sets. Two methods were used to determine positive amplification and their results are arranged in the same panel: real-time curves (top) and plots of RFU obtained from endpoint scanning (bottom). In the RFU plotting, the threshold (value shown above the line) for positive reactions and a separation from NTC reactions divide the plot area into 4 quadrants as illustrated in panel A: positives, negatives, false positives and NTC. A. Detection by SYTO-9. B. Single-plex DARQ LAMP. C. Duplex DARQ LAMP including both E1 and *ACTB* primers. D. *ACTB* detection from duplex DARQ LAMP. E. Summary of detection. Total number of positives, amplification speed (Cq) and correlation of results by real-time curve and end point scanning.

Although real-time monitoring of LAMP provides some level of quantitative information, heating to 60 °C with multiplex fluorescence requires a qPCR instrument or similar, and reactions must be incubated on the detection instrument for the entire reaction time. An alternative approach compatible with more instruments is to use endpoint fluorescence detection. We investigated the possibility of using end-point fluorescence scanning to detect signal in DARQ LAMP reactions. After incubating at 60 °C, the same plate used for real time monitoring was scanned for fluorescence levels in a BioTek Synergy Neo2 plate reader and the RFU values were plotted (Figure 2A-C, lower panel). The difference in signal levels between positive and negative LAMP or NTC reactions was easily distinguished using a threshold value set based on negative (background value), and calling positives accordingly matched 100% with that of real-time monitoring (Figure 2E). Similarly, the *ACTB* signal in duplex LAMP reactions was determined by scanning and matched the real time results (Figure 2D). This result demonstrates compatibility of endpoint, plate-reader measurements with multiplex DARQ LAMP, enabling use on a wider range of instrument types and increasing potential test throughput.

Next, we extended the number and range of targets for DARQ LAMP for the detection of a single target and an internal control in the presence of up to four sets of LAMP primers. Figure 3 shows results of 24 replicate LAMP reactions each with 50 copies of SARS-CoV-2 RNA and 8 without target using: E1 and *ACTB* primers (2-plex; Figure 3A); E1, *ACTB*, and IAV (3-plex, Figure 3B); or E1, *ACTB*, IAV and IBV (4 sets, Figure 3C). In all three cases, the SARS-CoV-2 RNA was successfully detected with equivalent sensitivity. We did observe a slowing of amplification in the presence of increasing primer amount, but sensitivity was not affected. Again, real-time results matched exactly with endpoint measurements on a plate reader (Figure 3A-C, lower half panel) for all primer combinations. The *ACTB* signal also appeared as expected and could be easily called as positive by the real-time data or endpoint fluorescence scanning in all reaction configurations (Figure 3D-E). A similar evaluation was performed with the IAV/Influenza A RNA and IBV/Influenza B RNA with equivalent performance observed with these targets and combinations of primers and templates (Supplementary Figures 1–2). These results demonstrate that DARQ LAMP detection sensitivity is not affected with up to 4 primer sets present in the reaction.

**Figure 3.**
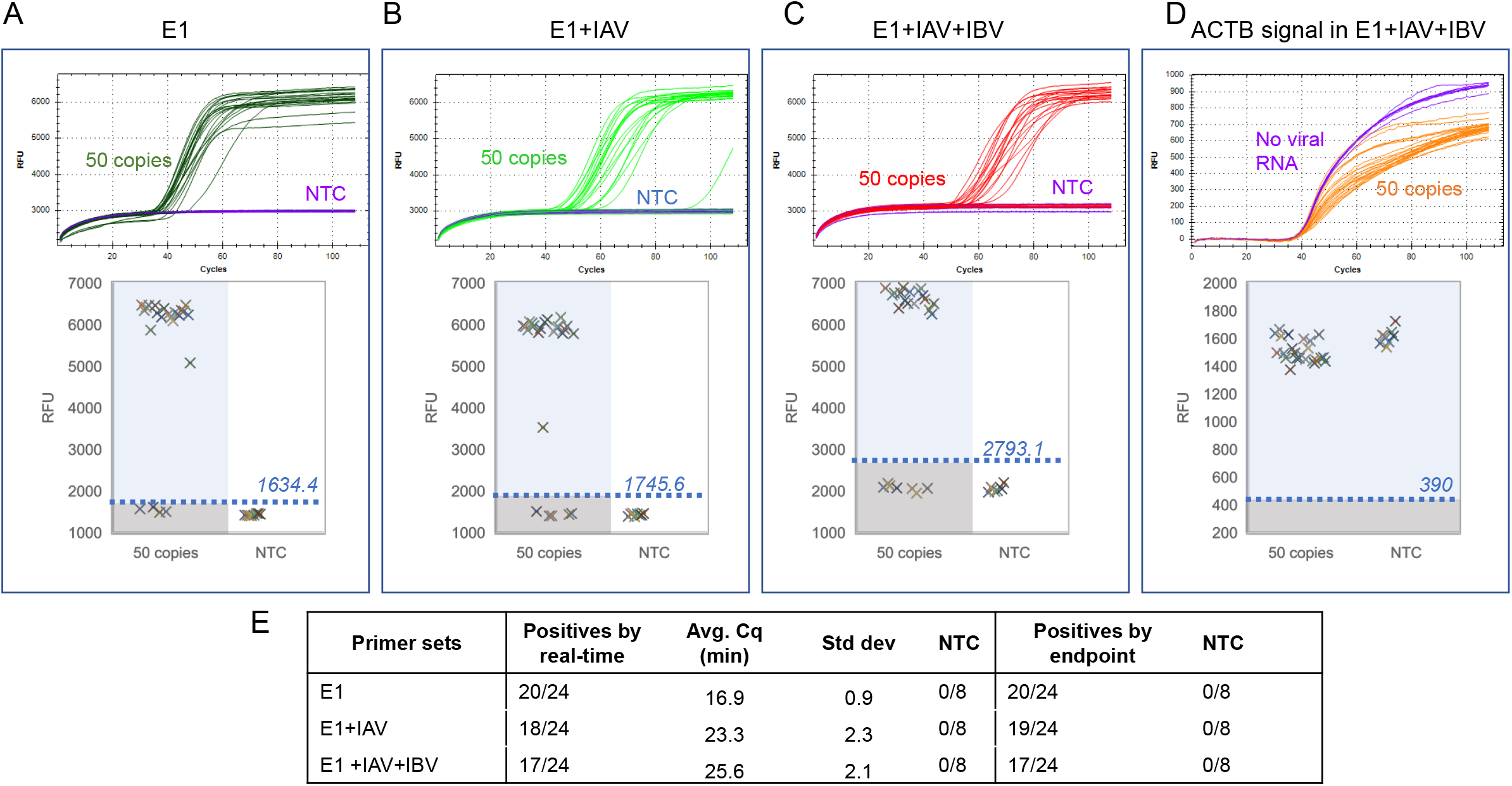
Sensitive detection of SARS-CoV-2 and an internal control in the presence multiple primer sets. Amplifications were performed with 24 repeats each containing 50 copies of SARS-CoV-2 RNA or 8 repeats as NTC using E1 and *ACTB* primer sets or with additional primer sets. A. Dual (E1 + *ACTB*) DARQ LAMP amplification. B. Dual (E1 + *ACTB*) DARQ LAMP amplification in the presence of a unrelated primer set (IAV and its QFIP:Fd duplex). C. Dual (E1 + *ACTB*) DARQ LAMP in the presence of 2 unrelated primer sets (IAV and IBV and their QFIP:Fd duplexes). D. *ACTB* detection in from conditions in (C). E. Summary of detection: total number of positives, amplification speed (Cq) and correlation of results by real-time curve and end point scanning.

With testing in real-world settings, the vast majority of samples will contain zero or one of the viral targets detectable in the multiplex assay. However, co-infections are possible and the assay must maintain performance in the presence of multiple target sequences. To study this scenario, we examined amplification of two DARQ LAMP targets and an internal control in the same reaction. We added two viral RNA targets to 24 DARQ LAMP reactions that all contained E1, IAV, IBV and *ACTB* primers (and Jurkat RNA for internal control) and monitored amplification by real-time and endpoint fluorescence scanning. In all three triplex combinations (SARS-CoV-2 + Influenza A + *ACTB*; SARS-CoV-2 + Influenza B + *ACTB*; Influenza A + Influenza B + *ACTB*), the positive rate of each target was equivalent to that of single-plex or duplex reactions shown above (Figure 3) indicating that sensitivity was unaffected. The majority of reactions showed detection of both viral targets with a minority showing only one of the targets (Figure 4). The shape of the real-time curves in these triple LAMP reactions (2 targets + *ACTB*) appeared slightly flatter and slower but distinction above background was easily seen, and with end-point fluorescence level (data not shown), positives could be differentiated from negatives in the same manner as described above.

**Figure 4.**
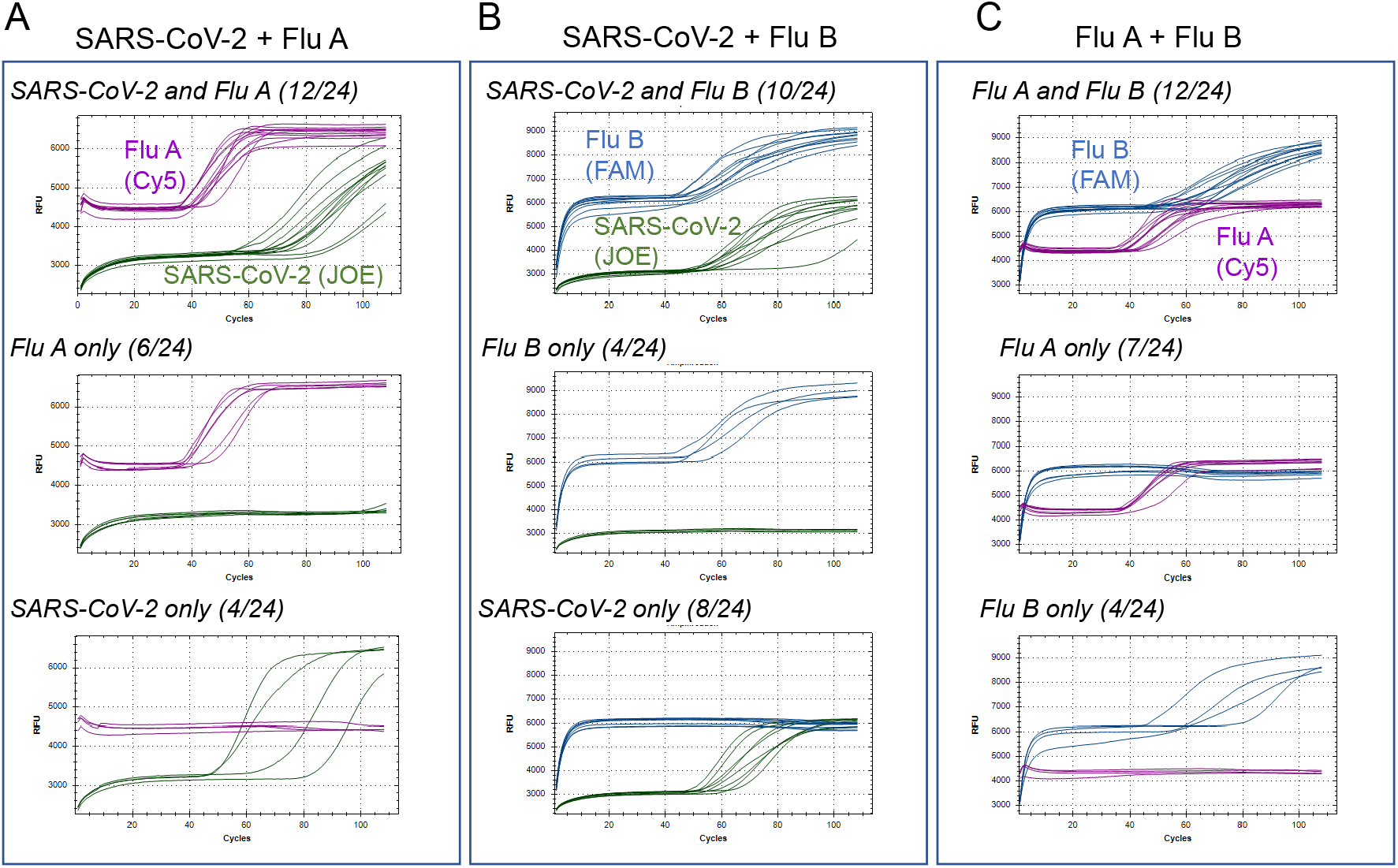
Simultaneous detection of 2 viral targets and an internal control. Reactions were performed in the presence of E1, IAV, IBV and *ACTB* primer sets and their QFIP:Fd duplexes with 24 repeats. The targets are pairs of SARS-CoV-2, Influenza A and Influenza B RNAs. For each target pair, the reactions that had both targets amplified are shown in the top, and reactions only that had one target amplified are shown in the middle and bottom. The fractional numbers in brackets indicate number of reactions belonging to each category. A. SARS-CoV-2, Influenza A, and Jurkat RNAs. B SARS-CoV-2, Influenza B, and Jurkat RNAs. C. Influenza A, Influenza B, and Jurkat RNAs.

Detection of multiple viral targets in a molecular diagnostic reaction is hardly our original idea and is widely used worldwide for multiplex PCR tests and syndromic panels. But as the COVID-19 pandemic continues into late 2020 and beyond, the need for more and varied testing remains an urgent priority worldwide. LAMP has found a use as a COVID testing modality due to its flexibility and simplicity, but as cold and flu season arrives more functionality will be needed in LAMP tests to keep pace with diagnostic testing requirements. While fieldable colorimetric LAMP is extremely useful for screening and surveillance, the ability to multiplex influenza and potentially other targets will supplement the supply-constrained RT-qPCR workflows. Here we demonstrate multiplex fluorescent DARQ LAMP with detection of three primary respiratory viral RNA targets and an internal control in a single reaction. While not as simple as visual colorimetric LAMP, the DARQ approach is rapid (<40 minutes) and is compatible with standard qPCR instruments. We also showed that positive signals can be reliably detected by endpoint scanning of DARQ fluorescence levels in a plate reader, and thus the reactions could be performed on heat blocks, regular PCR cyclers or even simple incubators followed with quick scanning. We hope by demonstrating this multiplex approach we can help enable broader usage of LAMP testing and further support the continuing need for diagnostics to combat the current COVID-19 pandemic. Ideally, as the power of molecular testing gains wider adoption we will continue to utilize these approaches, with LAMP and other diagnostic methods tracking and fighting the threats to public health that will inevitably arise in the future.

## Materials and Methods

DARQ LAMP primer sets include all conventional LAMP primers (F3, B3, FIP, BIP, LF and LB) and a duplex oligo consisting of the FIP modified at its 5′-end with a dark quencher (Q-FIP) annealed a complementary F1c oligo with 3′ fluorophore (Fd) [17] (Table 1). E1 LAMP primers targeting SARS-CoV-2 sequence (GenBank accession number MN908947) were from our previous screening [11]. IAV and IBV primer sets were designed and validated previously for targeting multiple Influenza A and B strains previously [25]. Oligos were synthesized at Integrated DNA Technologies (Coralville, IA) with standard desalting for conventional LAMP primers and HPLC purification for QFIP and Fd oligos. Synthetic SARS-CoV-2 RNA containing equal ratio of the viral genome regions was purchased from Twist Bioscience (Twist Synthetic SARS-CoV-2 RNA Control 2 #102024, MN908947.3). RNAs for Influenza A and B were purchased from ATCC: H1N1/2009pdm, ATCC VR-1737D; H1N1 A/PR/8/34, ATCC VR-1469DQ; H3N2 A/Virginia/ATCC6/2012; ATCC VR-1811D; H3N2 A/Aichi/2/68, ATCC VR-1680D; B/Wisconsin/1/2010 BX-41A, ATCC-1885DQ; B/Lee/40 ATCC VR-1535. Viral RNA was diluted to lower concentrations in 10 ng/μl Jurkat total RNA (Biochain) based on quantification provided by the manufacturers. For the 24 repeat reactions, the amount of RNA used was 50 copies of SARS-CoV-2 RNA, 1 ul of 1:1000 diluted Influenza A RNA and approximately 21 copies for Influenza B RNA. This amount of viral RNAs was sufficient for more than half but not all the 24 repeats to show positive amplification thus allowing detection of sensitivity change under different conditions.

**Table 1.**
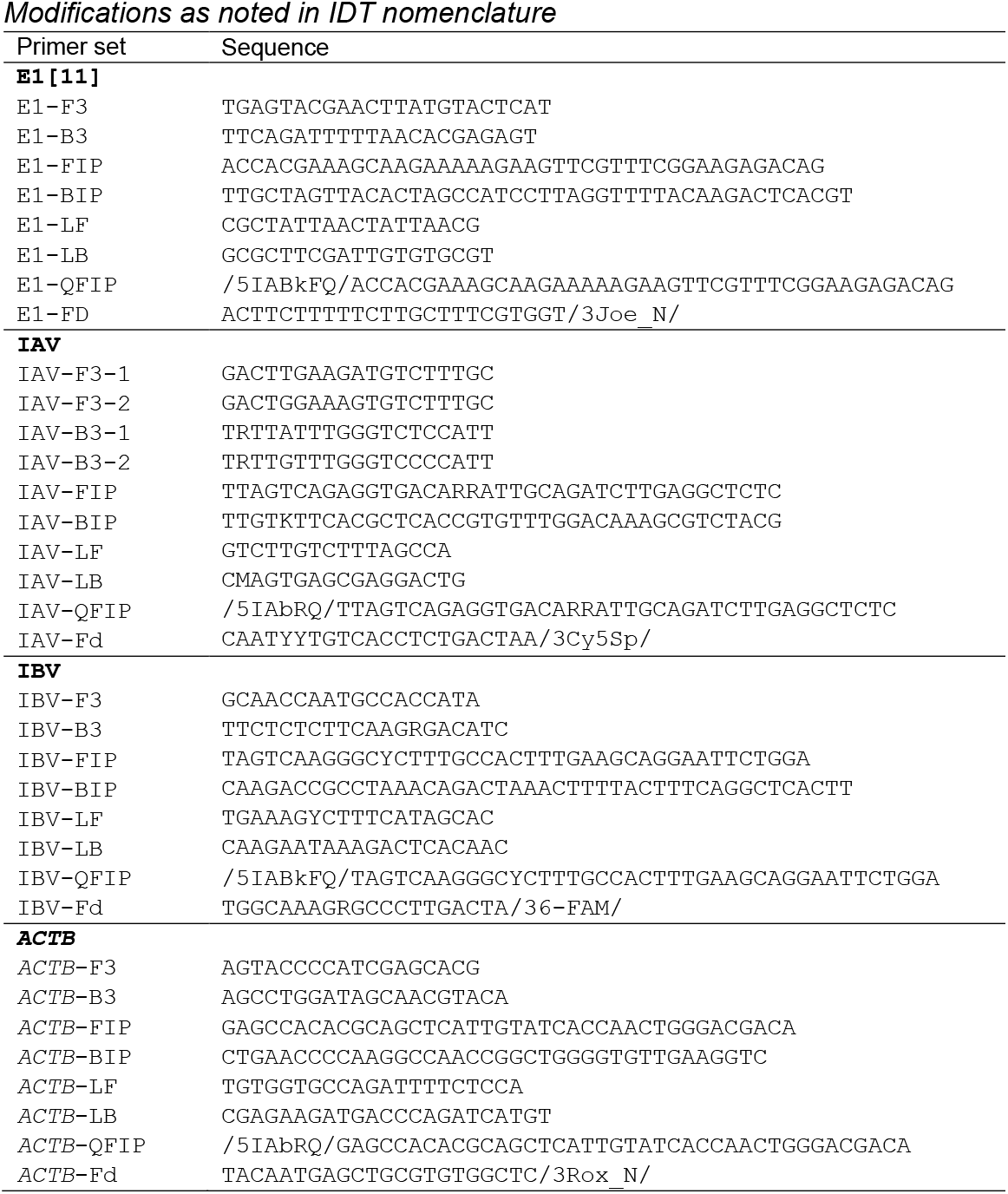
LAMP primer sequences (source)

All influenza primers were initially screened for performance using WarmStart® Colorimetric LAMP 2X Master Mix (DNA & RNA) (New England Biolabs, M1800) and with WarmStart LAMP Kit (DNA & RNA) (E1700) supplemented with 1 μM SYTO®-9 double-stranded DNA binding dye (Thermo Fisher S34854). DARQ LAMP reactions contained 1x E1700, with an additional 0.32 U/μL *Bst* 2.0 WarmStart DNA Polymerase (M0538), 40 mM guanidine hydrochloride (Sigma RDD001) and standard concentrations of LAMP primers (0.2 μM F3, 0.2 μM B3, 1.6 μM FIP, 1.6 μM BIP, 0.4 μM LoopF, 0.4 μM Loop B) supplemented with DARQ FIP duplex (0.22 μM QFIP: 0.18 μM Fd, pre-annealed as 55 μM QFIP: 45 μM Fd by heating to 95 °C and slowly cooling to room temperature). The concentration for *ACTB* primers was reduced to 0.25X the standard concentrations for LAMP primers (0.05 μM F3, 0.05 μM B3, 0.4 μM FIP, 0.4 μM BIP, 0.1 μM LoopF, 0.1 μM Loop B) with DARQ FIP duplex added as 0.066 μM QFIP and 0.054 μM Fd. The reactions were incubated at 60 °C in half-skirted plates (Bio-Rad HSP9601) on a real-time qPCR machine (Bio-Rad CFX96). Real time LAMP signal was acquired every 15 seconds for 108 cycles (total incubation time ∼40 min for single channel or 49 min for 4-channel acquisition). Completed LAMP reactions were then scanned on BioTek Synergy Neo2 microplate reader for fluorescence signal for 5-FAM (Excitation, 484/20, Emission 530/25, Signal Gain 75), HEX (524/20, 565/20, 75), 5-ROX (569/20, 615/25, 85) and Cy5 (640/20, 682/20, 75). The threshold for the positive signal was set as above the sum of average raw fluorescence units (RFU) of 8 NTC reactions plus 10x of their standard deviation.

## Data Availability

All data is in the manuscript

---

**Supplementary Figure 1.**
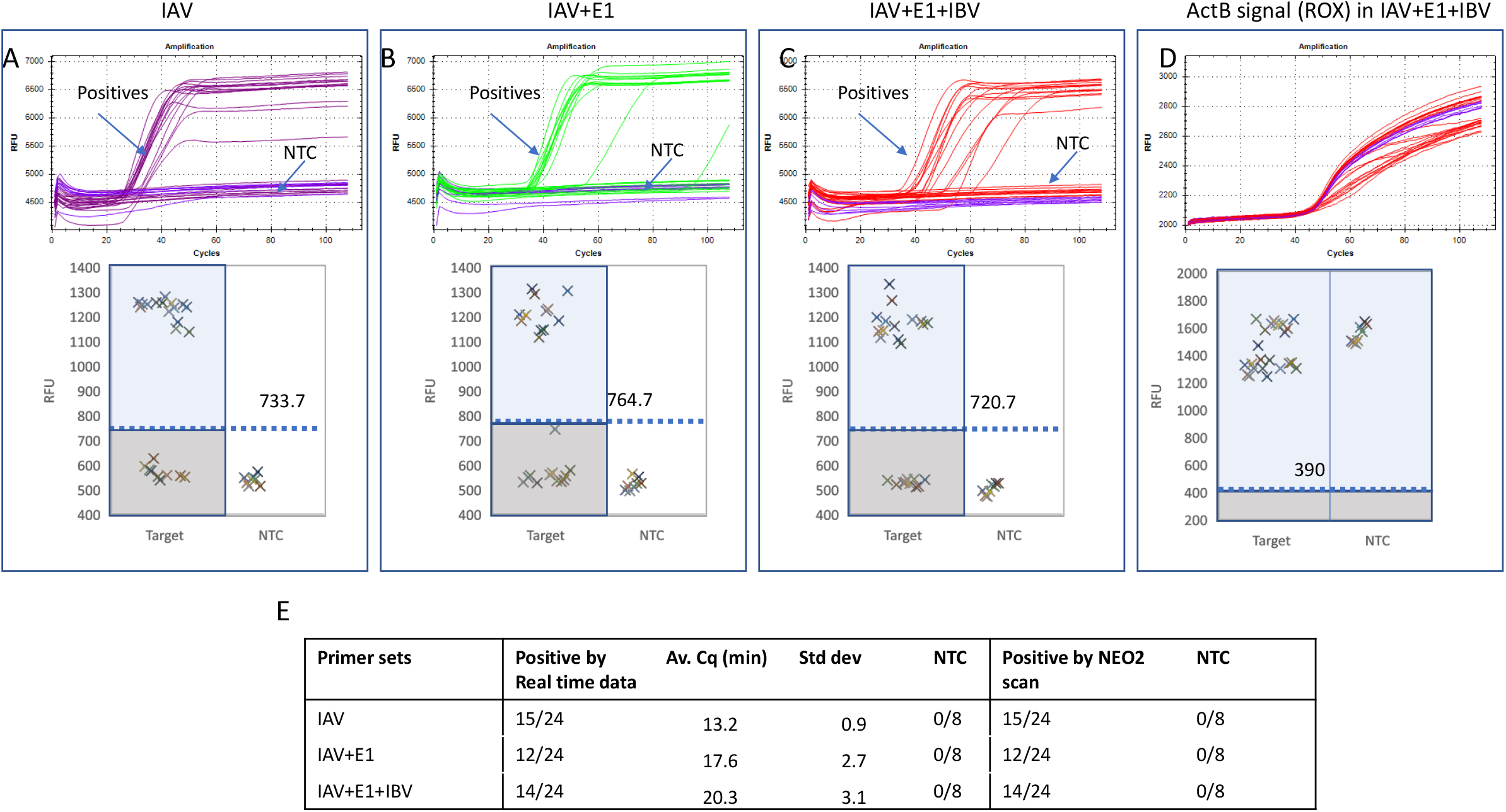
Detection of Influenza A RNA and an internal control in the presence multiple primer sets. Reactions were performed with 24 repeats each containing 1 μl of Flu A RNA diluted to 1/10000 or 8 repeats as NTC using both IAV and *ACTB* primer sets or with additional primer sets. The results were organized the same way as in Figure 2. A. Dual (IAV + *ACTB*) DARQ LAMP amplification. B. Dual (IAV + *ACTB*) DARQ LAMP amplification in the presence of a unrelated primer set E1 and its QFIP:Fd duplex. C. Dual (IAV + *ACTB*) DARQ LAMP in the presence of 2 unrelated primer sets E1 and IBV and their QFIP:Fd duplexes. D. *ACTB* detection in C. E. Summary of detection: total number of positives, amplification speed (Cq) and correlation of results by real-time curve and end point scanning.

**Supplementary Figure 2.**
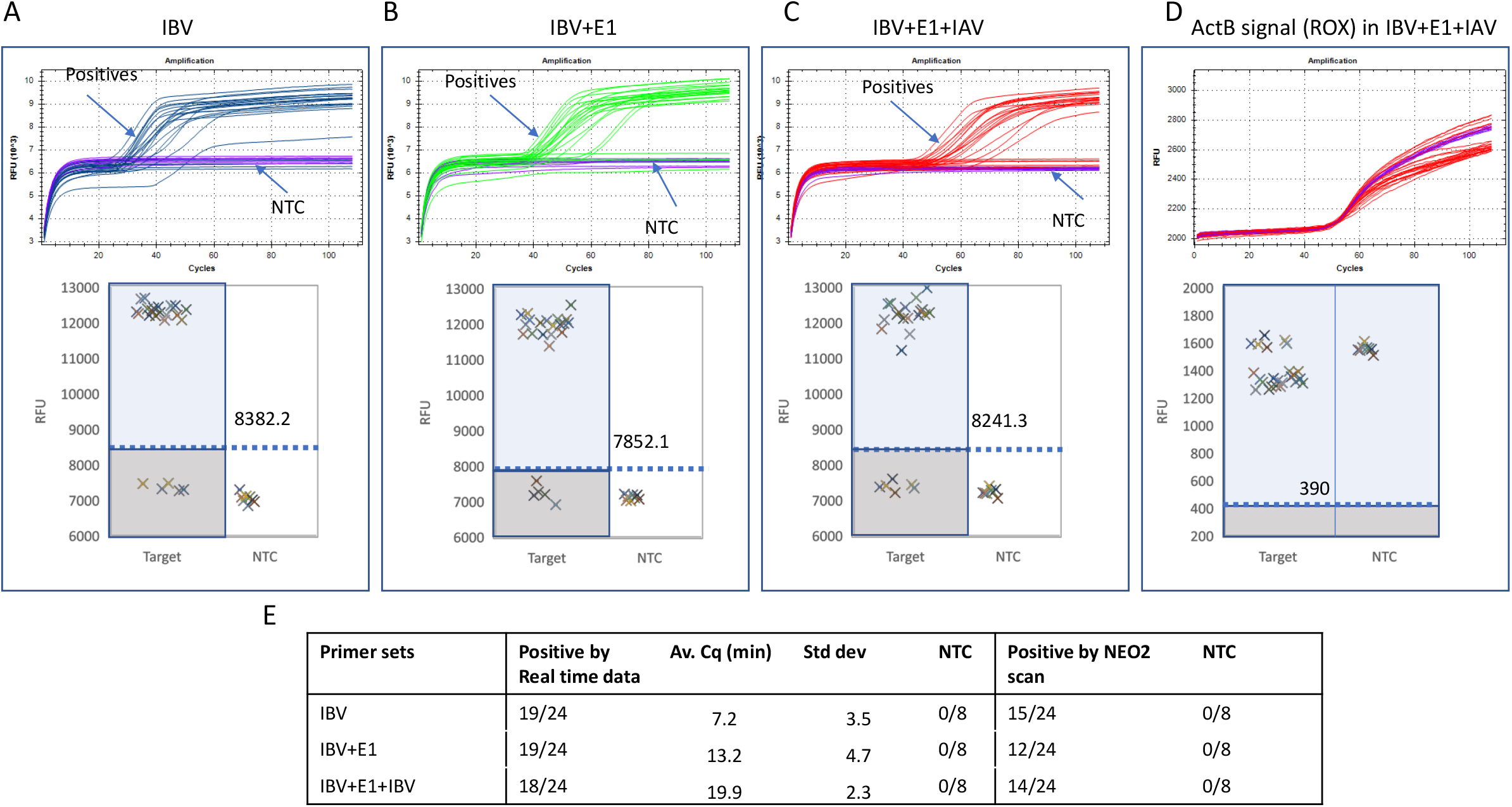
Detection of Influenza B RNA and an internal control in the presence multiple primer sets. Reactions were performed with 24 repeats each containing ∼21 copies of Influenza B RNA or 8 repeats as NTC using IBV and *ACTB* primer sets or with additional primer sets. The results were organized the same way as in Figure 2. A. Dual (IBV + *ACTB*) DARQ LAMP amplification. B. Dual (IBV + *ACTB*) DARQ LAMP amplification in the presence of a unrelated primer set E1 and its QFIP:Fd duplex. C. Dual (IBV + *ACTB*) DARQ LAMP in the presence of 2 unrelated primer sets E1 and IAV and their QFIP:Fd duplexes. D. *ACTB* detection in C. E. Summary of detection: total number of positives, amplification speed (Cq) and correlation of results by real-time curve and endpoint scanning.

